# Non-autoimmune diabetes in young people from Assam, India: the PHENOEINDY-2 study

**DOI:** 10.1101/2024.02.02.24302249

**Authors:** Anupam Dutta, Pranjal K. Dutta, Sreemanta M. Baruah, Prasanta Dihingia, Arpita Ray, Dattatrey S. Bhat, Sonali W. Patki, Pradeep Tiwari, Madhura Deshmukh, Rucha Wagh, Rubina Mulchandani, Tanica Lyngdoh, Sanjeeb Kakati, Chittaranjan S. Yajnik

**Affiliations:** Department of Medicine, Assam Medical College and Hospital, Dibrugarh, India; Diabetes Unit, Kamalnayan Bajaj Diabetology Research Centre, King Edward Memorial Hospital and Research Centre, Pune, India; Hubert Department of Global Health, Rollins School of Public Health, Emory University, Atlanta, GA, USA; Dr D.Y. Patil Medical College, Hospital and Research Centre, Dr D.Y. Patil Vidyapeeth, Pune, India; Symbiosis School of Biological Sciences, Symbiosis International (Deemed University), Pune, India; Indian Institute of Public Health Delhi, Public Health Foundation of India, Gurgaon, India; Division of Reproductive, Child Health and Nutrition (RCN), Indian Council of Medical Research, New Delhi, India; Faculty of Biological Sciences, Academy of Scientific & Innovative Research (AcSIR), Council of Scientific and Industrial Research-Human Resource Development Centre (CSIR-HRDC), Ghaziabad, Uttar Pradesh, India

**Author notes:** **Corresponding authors:** Chittaranjan Yajnik, Sanjeeb Kakati. **Social media post** Non-autoimmune diabetes in undernourished young people in NE India reveals intricacies of diabetes phenotypes in rapidly transiting populations. Rural lean people show reduced insulin secretion; urbanised overweight people show insulin resistance Received: 13 March 2025/Accepted: 28 April 2025.

**Keywords:** Adiposity, Assam, Beta cell deficiency, Body roundness index, India, Lean, Truncal adiposity, Young-onset diabetes

## Abstract

**Aims/hypothesis:** In the Western world, non-autoimmune diabetes in the young is believed to be driven by overweight/obesity and insulin resistance. However, it is increasingly being reported in undernourished people in low– and middle-income countries, including India. We hypothesised that these patients would show markers of chronic undernutrition and a ‘thin–fat’ phenotype and be predominantly beta cell-deficient.

**Methods:** We studied young patients (clinically diagnosed with type 2 diabetes at < 40 years) who attended the outpatient department of Assam Medical College and Hospital, Dibrugarh (in North-East India). We measured weight, height, waist and hip circumference, haemoglobin, fasting glucose, HbA_1c_, lipid, GADA and C-peptide levels, and body fat percentage (adiposity, assessed using dual-energy x-ray absorptiometry), and calculated BMI (kg/m^2^), body roundness index and HOMA indices. Volunteers from similar socioeconomic background with normal glucose tolerance (measured by 75 g OGTT) were assessed as control participants. We also compared the anthropometric characteristics and body composition of our participants with those of non-Hispanic white Americans from the NHANES study.

**Results:** The study included 252 control participants (136 male participants, median age 30 years, BMI 23.0 kg/m^2^) and 240 GADA-negative young patients with diabetes (155 male participants, age 36 years, BMI 23.0 kg/m^2^). The majority of study participants came from a relatively impoverished population of tea garden workers (‘tribals’). Of the patients with diabetes, 28% had stunted growth (male <161.2 cm, female <149.8 cm), 27% were anaemic, 68% were lean (BMI <25 kg/m^2^, including 14% who were underweight [BMI <18.5 kg/m^2^]) and 32% were overweight/obese (BMI ≥25 kg/m^2^). When assessed using dual-energy x-ray absorptiometry, 61% of control participants and 53% of patients had adiposity (body fat percentage >25% in male participants or >35% in female participants). Compared with a contemporary non-Hispanic white American population, Assamese control participants and diabetic patients had higher WHR, body roundness index, and total and truncal adiposity (assessed using dual-energy x-ray absorptiometry) across the range of BMI, thus conforming to the description of the ‘thin–fat’ phenotype. The diabetic patients were severely beta cell-deficient (median HOMA-B 25.7) and only moderately insulin-resistant (median HOMA-S 103) with higher triacylglycerol and lower HDL-cholesterol concentrations than control participants. Underweight patients (<18.5 kg/m^2^) were the most hyperglycaemic (based on fasting plasma glucose and HbA_1c_), and were severely beta cell-deficient but insulin-sensitive. As previously reported, two-thirds of these patients belonged to the severely insulin-deficient diabetes (SIDD) cluster according to the Swedish diabetes subgroup classification.

**Conclusions/interpretation:** Diabetes in the young people of this impoverished population is heterogeneous, but the majority of patients are not overweight/obese or insulin-resistant. Overall, these participants conform to the thin–fat phenotype, and their diabetes is predominantly driven by beta cell deficiency. The sociodemographic history and physical characteristics of this population suggest a role for multigenerational undernutrition in the aetiology of non-autoimmune diabetes in these young patients from Assam.

## Introduction

Non-autoimmune diabetes is considered to be a metabolic endocrine disorder associated with ageing, overnutrition/obesity and a combination of insulin resistance and reduced insulin secretion [1]. Usually, it is called type 2 diabetes. The diabetes epidemic has rapidly grown across the world, especially in low– and middle-income countries, including India, which is now almost considered the world capital of diabetes [2, 3]. Type 2 diabetes is diagnosed at a much younger age and a lower BMI in India (and in other low– and middle-income countries) compared with Europe [4]. This may be partly genetically determined [5], but is more likely due to the *‘*thin–fat’ (thrifty) phenotype commonly seen in people of South Asian/Indian ancestry [6]. This phenotype originates in utero due to the influence of maternal undernutrition and continues through childhood and puberty into adult life [7–11]. Most research papers describing the characteristics of Indian patients with diabetes include participants from metropolitan cities and stress the role of an obesogenic lifestyle and associated insulin resistance [12]. The INDIAB study described the heterogeneity of the phenotype of type 2 diabetes across India [13] and the GBD-India study highlighted that the most rapid rise in the prevalence of diabetes in the last 25 years has occurred in chronically deprived parts of the country such as Uttar Pradesh, Chhattisgarh and Odisha [14]. There have been reports of lean type 2 diabetes patients from rural impoverished populations in the states of Odisha [15], Assam [16] and Tamil Nadu [17]. Such patients have also been reported in other low– and middle-income countries, including countries in Africa (Uganda and Ethiopia) [18–20], South-East Asia [21, 22] and the Caribbean (Jamaica) [23,24]. Increasing food insecurity in many parts of the world due to climate change, famines, floods, wars and migrations will probably contribute to a higher prevalence of such a phenotype across the world in the future.

The north-east of India is unique in its ethnic diversity and history of hardships due to ecological stresses, colonial rule, exclusion and political unrest, dating back hundreds of years [25]. The state of Assam has the lowest life expectancy at birth in the country (66.2 years for Assam vs 69.0 years for India) [26], a low per capita income (US$1368 per annum vs the Indian average of US$2191 per annum), and a low BMI (second only to the state of Jharkhand) [14], but showed an increase in the prevalence of diabetes in adults from 5.5% in 1990 to 7.7% in 2016 [13]. The Comprehensive National Nutrition Survey showed that the prevalence of prediabetes in children and adolescents (fasting plasma glucose 5.5–6.9 mmol/l and HbA_1c_ 39–46 mmol/mol [5.7– 6.4%]) is unexpectedly high in relatively undernourished populations, including in some states from the north-east of India [27]. The Young Diabetes Registry of India (which includes patients who are diagnosed before 25 years of age) reported a higher proportion of young-onset type 2 diabetes in Assam compared with other states [28].

We studied young patients with non-autoimmune diabetes (diagnosed at < 40 years of age) who attended the outpatient clinic of Assam Medical College and Hospital. We reasoned that studying their physical and metabolic endocrine characteristics would provide insight into the aetiology and pathophysiology of diabetes in impoverished young patients, and guide appropriate prevention and treatment strategies for such populations who may otherwise be treated using Western guidelines.

## Methods

### Study design and setting

This is a hospital-based case–control study. Patients were recruited from outpatients attending the Department of Medicine, Assam Medical College and Hospital (AMCH), Dibrugarh, India. AMCH is the first medical college in North-East India and was established in 1947 (formerly Berry White Medical School from 1901 to 1947). It is a tertiary medical referral centre for upper Assam and areas in neighbouring states. It is a lifeline for disadvantaged people in the region, including Indigenous and ‘tribal’ tea garden workers (tea tribes are the people who were forced to migrate by the British to work in tea gardens; see ESM Background Text).

### Sampling method and sample size

We used purposive sampling to select patients. The proposed sample size of 500 (patients plus control participants), to be recruited over a period of 3 years, was based on the annual footfall of approximately 1000 in the diabetes outpatient department of AMCH over the last 5 years and an estimated proportion of approximately 10% of these patients being diagnosed before 40 years of age.

### Study population

The patients included were those with diabetes who attended the outpatient department of AMCH, diagnosed below the age of 40 years, and clinically classified as having type 2 diabetes. Diabetes was diagnosed according to the WHO/IDF criteria [29]: fasting plasma glucose ≥ 7.0 mmol/l, postprandial glucose ≥ 11.1 mmol/l, HbA_1c_ ≥ 48 mmol/mol (6.5%). They were enrolled in the PHEnotypingNOrthEastINDianYoung type 2 diabetes (PHENOEINDY-2) study if they were willing to participate in physical measurements and blood tests. We excluded patients who were clinically diagnosed with type 1 diabetes (indicated either by GADA positivity or by all of the following criteria: diagnosed before 20 years of age, history of diabetic ketoacidosis, lifelong insulin treatment), had fibrocalculous pancreatic diabetes (clinical symptoms, and x-ray or sonographic evidence of chronic pancreatitis), or were clinically suspected to have MODY. We also excluded those with recent hospital admission (in the last 6 weeks) for treatment of a medical condition. Control participants included acquaintances, friends or colleagues of the patients from similar geographic and socioeconomic backgrounds. They were included in the study if a 75 g OGTT showed normal glucose tolerance (NGT) according to the WHO/IDF criteria. Consenting participants were recruited serially between May 2017 and May 2020. These participants are representative of the outpatient attendees at AMCH.

### Study procedures and measurements

Participants were instructed to eat their normal dinner the previous evening, and to visit the AMCH Diabetes Research Unit in the morning after an overnight fast. Investigations included a health questionnaire, physical examination, anthropometric and body composition measurements, and blood tests. All measurements were made by trained and certified staff. The interviewer-assisted questionnaire collected information on sociodemographic characteristics, ethnicity, substance use and family history of diabetes. Smoking was defined as present if the participant reported being a current smoker, and alcohol consumption was defined as consumption three or more times a week. Socioeconomic status was assessed by calculating the standard of living index (SLI) score described previously [30].

Weight was measured using a calibrated digital scale (EURO Weight Machine, Coimbatore, Tamil Nadu, India), and height was measured using a fixed stadiometer (SECA model 755, Hamburg, Germany). Waist and hip circumferences were measured using a fibreglass measuring tape (SECA model 203). Skinfold thicknesses (biceps, triceps, subscapular and suprailiac) were measured using skin calipers (CMS Instruments, Harpenden, UK). The mean of three readings at each site was used to calculate the sum of the four skinfold thicknesses. Body composition was measured by dual-energy x-ray absorptiometry (DEXA) using a Prodigy model (Lunar, Madison, WI, USA), according to the manufacturer’s guidelines. The accompanying software reports body composition as the mass and proportions (%) of total and regional fat, lean matter (muscle and organs), bone and water.

Stunting refers to a height <161.2 cm for men and <149.8 cm for women (corresponding to 2 SD below the height for adults in WHO standards) [31]. BMI was categorised as underweight (<18.5 kg/m^2^), normal (18.5–24.9 kg/m^2^), overweight (25.0–29.9 kg/m^2^) and obese (≥30.0 kg/m^2^). ‘Lean’ refers to people with a BMI <25 kg/m^2^. Central obesity was assessed on the basis of waist circumference (≥85 cm in men or ≥80 cm in women) or WHR (≥0.90 in men or ≥0.85 in women). The body roundness index (BRI) was calculated as described by Zhang et al [32]. Adiposity was defined as described by the American Association of Clinical Cardiology (body fat percentage >25% in men or >35% in women as measured using DEXA) [33]. Truncal adiposity (measured using DEXA) was calculated as ratio of truncal to total fat mass (expressed as a percentage).

We used publicly available data for anthropometric characteristics and DEXA body composition from the NHANES cohort of non-Hispanic white Americans aged 18–50 years for comparison with our cohort, across the BMI range for Assamese participants in our cohort (15–33 kg/m^2^) [34].

A fasting venous blood sample was collected. In control participants, a 75 g anhydrous OGTT was performed, and glucose tolerance was classified as per WHO/IDF criteria [29]; participants with NGT were used to obtain data for comparison with the diabetic patients. Clinical laboratory assays were performed at the Central Laboratory of AMCH. Plasma glucose, total cholesterol, HDL-cholesterol and triacylglycerols were measured using standard enzyme assays on a Vitros 5600 auto-analyser (QuidelOrtho, San Diego, CA, USA). HbA_1c_ was measured by HPLC using a VARIANT II haemoglobin testing system (Bio-Rad, Hercules, CA, USA).

The following measurements were performed at the Diabetes Unit, King Edward Memorial Hospital, Pune, India using blood samples transported from AMCH. Plasma C-peptide was measured using an ELISA kit (Diagnostics Biochem Canada, London, ON, Canada) calibrated against WHO-IS 84/510 [35]. GADA was measured in the plasma of diabetic patients by direct electrochemiluminescence immunoassay (RSR, Cardiff, UK), those with values above 5 U/ml were categorised as having autoimmune diabetes. The plasma immunoreactive trypsinogen concentration was measured by ELISA (MyBioSource, San Diego, CA, USA). HOMA-B and HOMA-S were calculated using fasting glucose and C-peptide concentrations using the HOMA2 calculator [36]. This calculator excludes those with a fasting glucose concentration of more than 25 mmol/l.

### Statistical analysis

Values for continuous variables are presented as medians and IQR, and those for categorical variables are presented as percentages. Differences between groups were tested using the Wilcoxon rank sum test for continuous data or Fisher’s exact test for categorical data. We also used ANCOVA, adjusting for appropriate confounders based on prior knowledge and scientific relevance, to test differences and trends across groups. As this is an exploratory analysis in a rarely studied group of patients, a *p* value of 5% was considered statistically significant without using correction for multiple comparisons (nominal significance).

Our primary analysis was to describe the sociodemographic, physical, metabolic and endocrine characteristics of young non-autoimmune patients with diabetes, and to compare them with NGT control participants. Body composition, metabolic markers and endocrine function differ by sex, and male and female participants were therefore analysed separately. Given our interest in undernutrition-related diabetes, and the strong association of nutrition with BMI, we also compared patients and control participants by BMI categories: lean (BMI <25 kg/m^2^, subdivided into underweight [BMI <18.5 kg/m^2^] and normal [BMI 18.5–24.9 kg/m^2^]) and overweight/obese (BMI ≥ 25 kg/m^2^).

The original thin–fat phenotype of Indians was described based on a comparison with data from white people of European descent [8,9,37]. We compared the waist circumference, WHR, BRI, total and truncal adiposity for all our participants with the publicly available NHANES cohort data for contemporary non-Hispanic white Americans.

Sensitivity analysis included comparing diabetic patients and control participants matched by age using the ‘matchit’ function in R version 4.1.3 because of the significant difference in the age of patients and control participants. We also analysed data by the duration of diabetes, to compare characteristics of newly diagnosed patients (< 1 year since diagnosis) with those of control participants to minimise the effect of long-term gluco– and lipotoxicity.

Statistical analysis was performed using R software, version 4.1.3 (The R Project for Statistical Computing, Vienna, Austria).

### Ethical considerations

This study was approved by the Institutional Ethics Committee, Assam Medical College (AMC/EC/932, 21 March 2017). Written informed consent was obtained from all participants before data collection. Patients were provided with clinically relevant results and appropriately advised.

## Results

Between May 2017 and May 2020, we approached people with diabetes attending the outpatient department of AMCH to request their participation in the PHENOEINDY-2 study, based on the inclusion and exclusion criteria. Two hundred and seventy-nine patients with diabetes diagnosed before 40 years of age agreed, and 246 participated. Of these, six patients were positive for GADA and were excluded from further analysis. Thus, the final analysis was performed on 240 non-autoimmune young patients with diabetes and 252 control participants with NGT. The selection and flow of the study participants is presented in the flowchart shown in electronic supplementary material [ESM] Fig. 1. The percentages of missing data ranged from 1–3% for demographic and anthropometric characteristics, 5–9% for laboratory tests, and 12% for body composition measurements (using DEXA). Four diabetic patients with a fasting plasma glucose concentration >25 mmol/l were excluded from the HOMA calculation because of the limit imposed by the HOMA2 calculator.

**Fig. 1.**
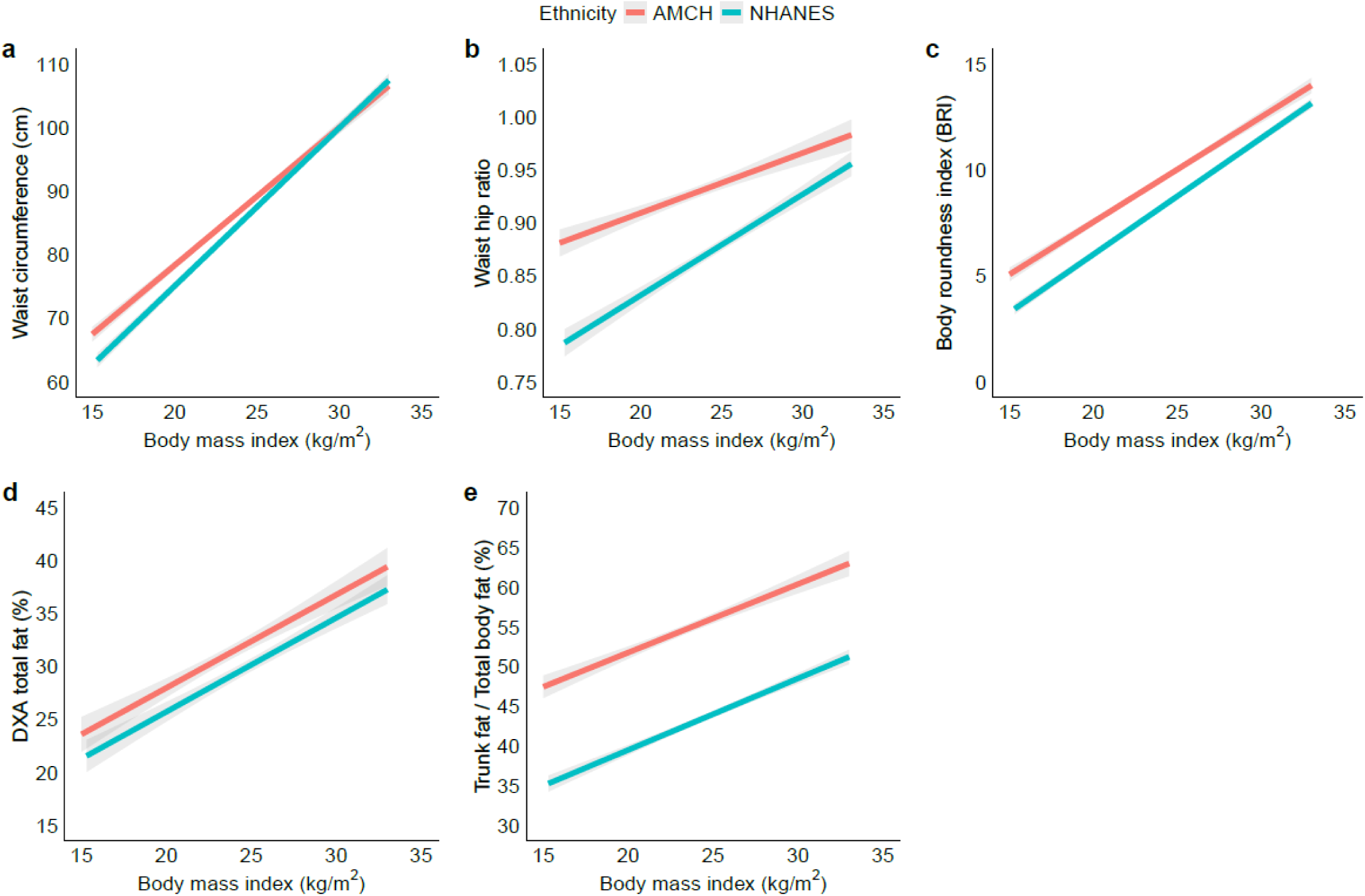
Comparison of waist circumference, WHR, BRI and total and truncal adiposity measured using DEXA in participants from AMCH (red line) and non-Hispanic white Americans (NHANES 2017–2020) (blue line). The Assamese population had higher indices across the range of BMI

### Sociodemographic and clinical characteristics

ESM Table 1 shows sociodemographic profile of the participants. Most were Assamese-speaking people from rural areas, and there was a balanced representation of religions (Hindu, Muslim or other) and ethnic groups. Both patients and control participants came from relatively low socioeconomic groups (low SLI score), and the majority were tribal tea garden workers.

**Table 1.**
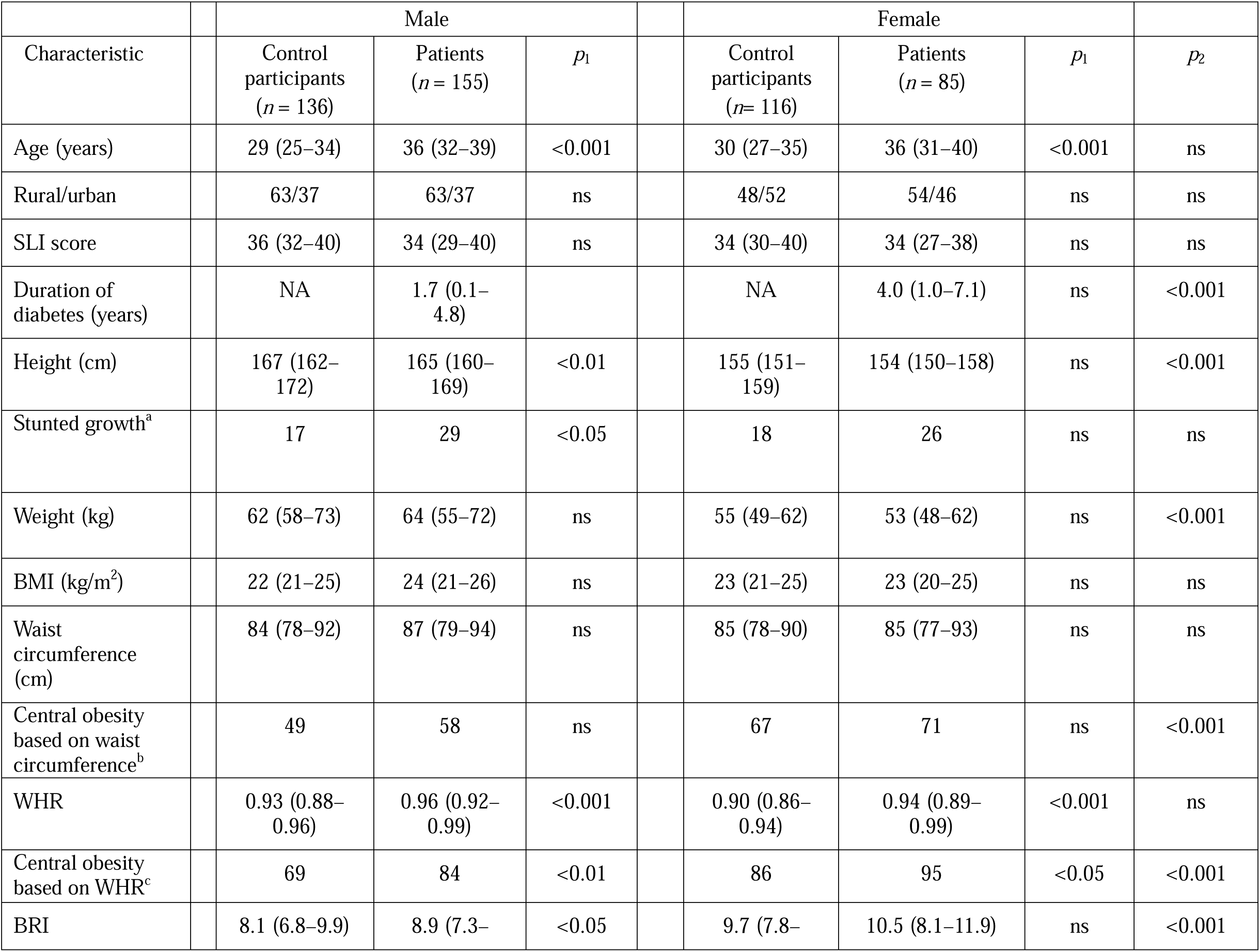
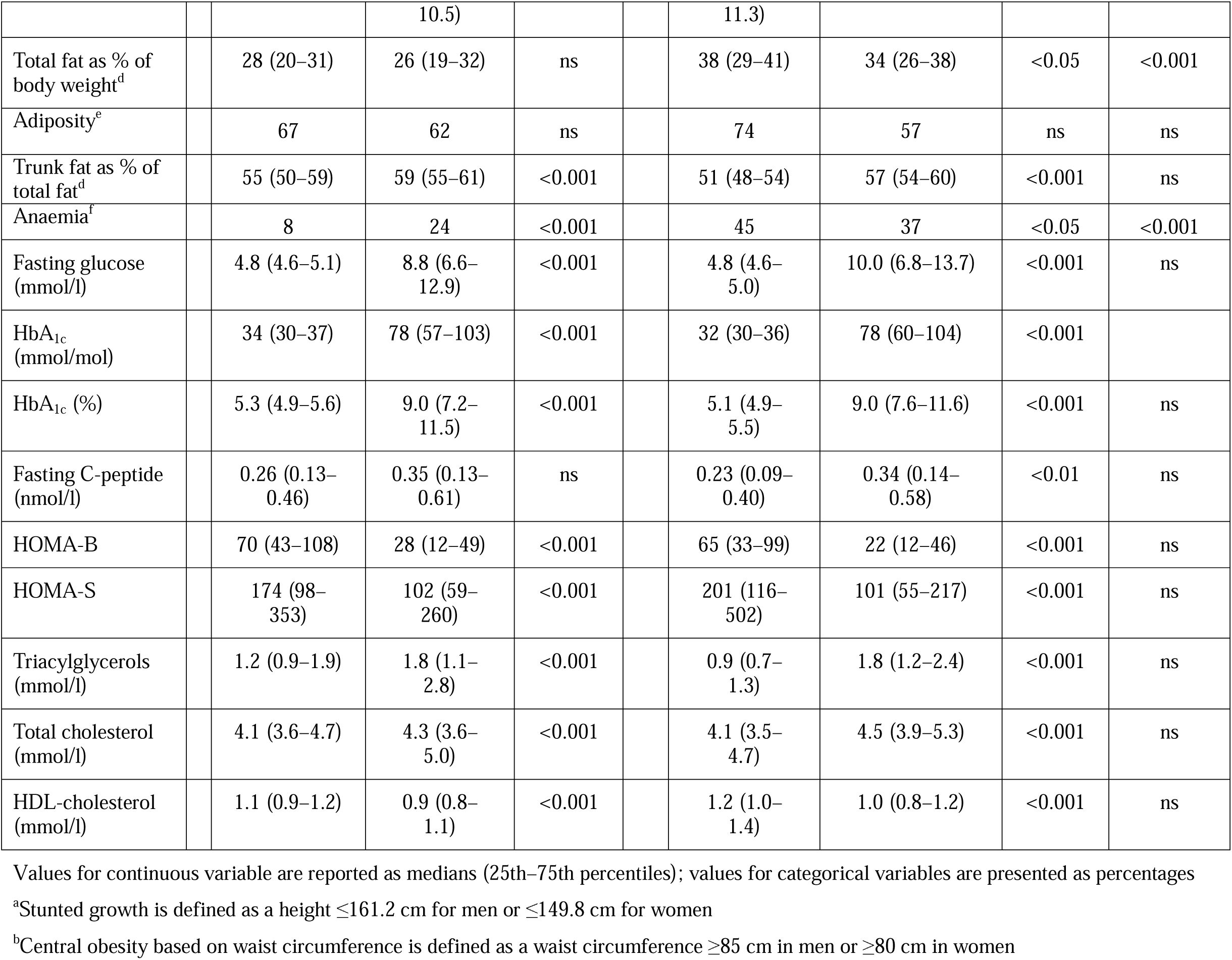
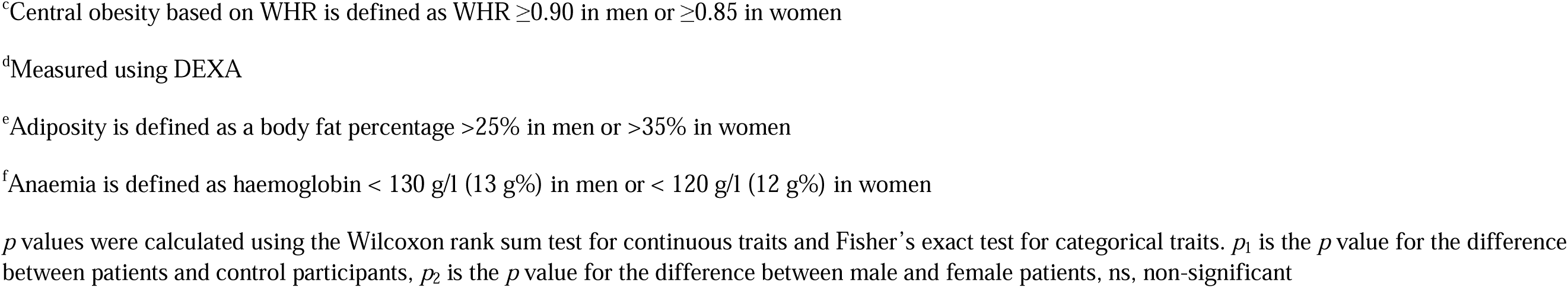
Characteristics of young patients with diabetes and control participants attending the outpatient department of AMCH.

The median age at diagnosis of diabetes was 36 years, and patients were older than control participants at the time of study. Patients were less educated, and consumed alcohol more frequently (ESM Table 1). Anaemia was common in this population (25% in control participants and 29% in patients), 29% participants had microcytosis and 27% had hypochromia. The median duration since diagnosis of diabetes was 2 years for male participants (IQR 0–5 years) and 4 years for female participants (IQR 1–7 years) (Table 1). A family history of diabetes in first-degree relatives was more prevalent in patients compared with control participants (55% vs 29%). Twenty patients (8%) received insulin treatment and 149 (62%) were taking oral glucose-lowering agents; five patients (2%) were taking anti-hypertensive drugs and two (1%) were taking cholesterol-lowering drugs.

### Anthropometry and body composition

As a group, the participants in our study (patients plus control participants) were of short stature (median height for male participants was 166 cm and that for female participants was 154 cm), and 17% of control participants and 28% of patients had stunted growth, according to WHO criteria. The median BMI was 23 kg/m^2^: 68% of patients were lean (BMI <25 kg/m^2^), including 14% who were underweight (BMI <18.5 kg/m^2^), and 32% were overweight/obese (BMI >25 kg/m^2^), including 5% who were obese (BMI >30 kg/m^2^).

Based on waist circumference, 56% of control participants and 62% of patients had central obesity; based on WHR, the proportions were 75% and 87%, respectively (Table 1). We also assessed central obesity on the basis of the recently described BRI [32], and found that patients had a higher BRI compared with control participants.

DEXA measurements showed that 64% of male participants and 66% of female participants had adiposity (based on body fat percentage) (Table 1). Male patients had similar DEXA-measured adiposity to that in male control participants, while female patients had significantly lower adiposity compared with control participants (ESM Table 2). We investigated truncal adiposity by calculating DEXA-measured truncal/total fat mass. Patients had higher truncal adiposity compared with control participants (Table 1). Truncal adiposity increased with increasing BMI; however, even in underweight patients, truncal fat accounted for 48% of total body fat (Table 2). To investigate the thin–fat paradigm in Assamese Indians, we compared our data with those for the contemporary non-Hispanic white American population in the NHANES cohort [34]. Our participants (both with and without diabetes) had similar waist circumference but higher WHR, higher BRI and higher total and truncal adiposity across the range of BMI, supporting the thin– fat phenotype of Assamese Indians (Fig. 1) [8]. Similar patterns were observed for male participants and female participants when analysed separately (ESM Fig. 2).

**Fig. 2.**
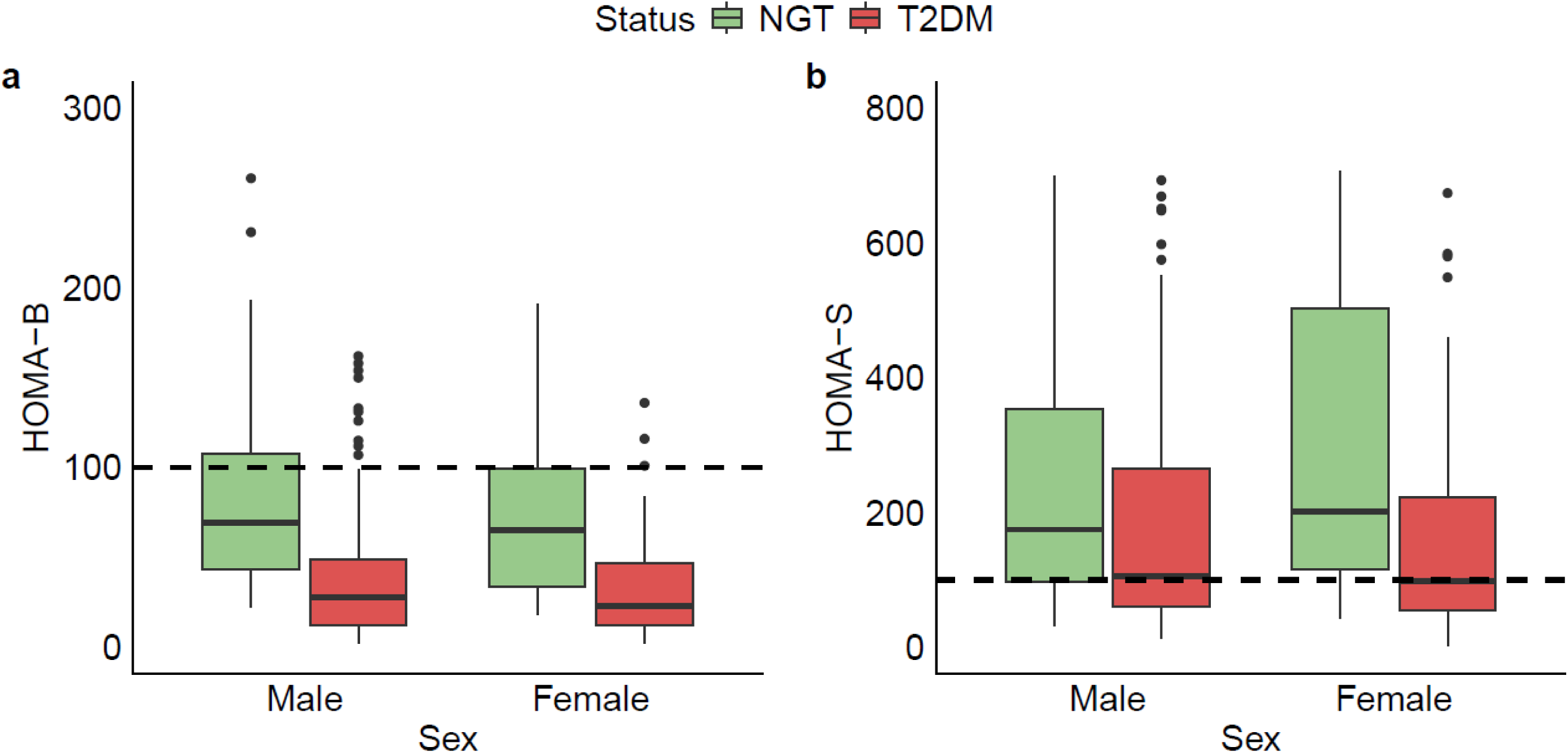
Comparison of (**a**) beta cell function (HOMA-B) and (**b**) insulin sensitivity (HOMA-S) between control participants (green bars) and young patients with type 2 diabetes (red bars). The dotted line at 100 represents control values from young UK patients [32]

**Table 2.**
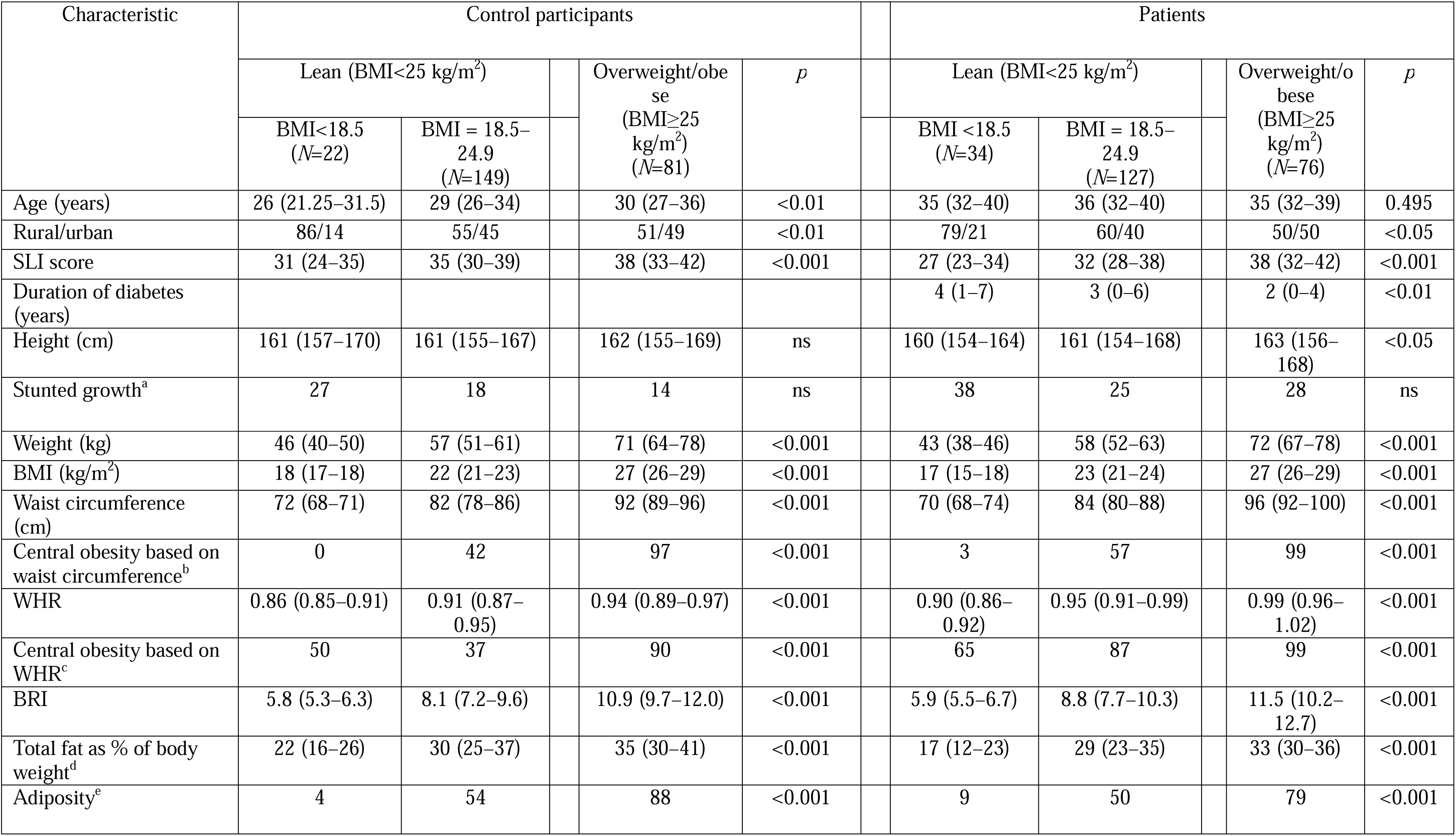
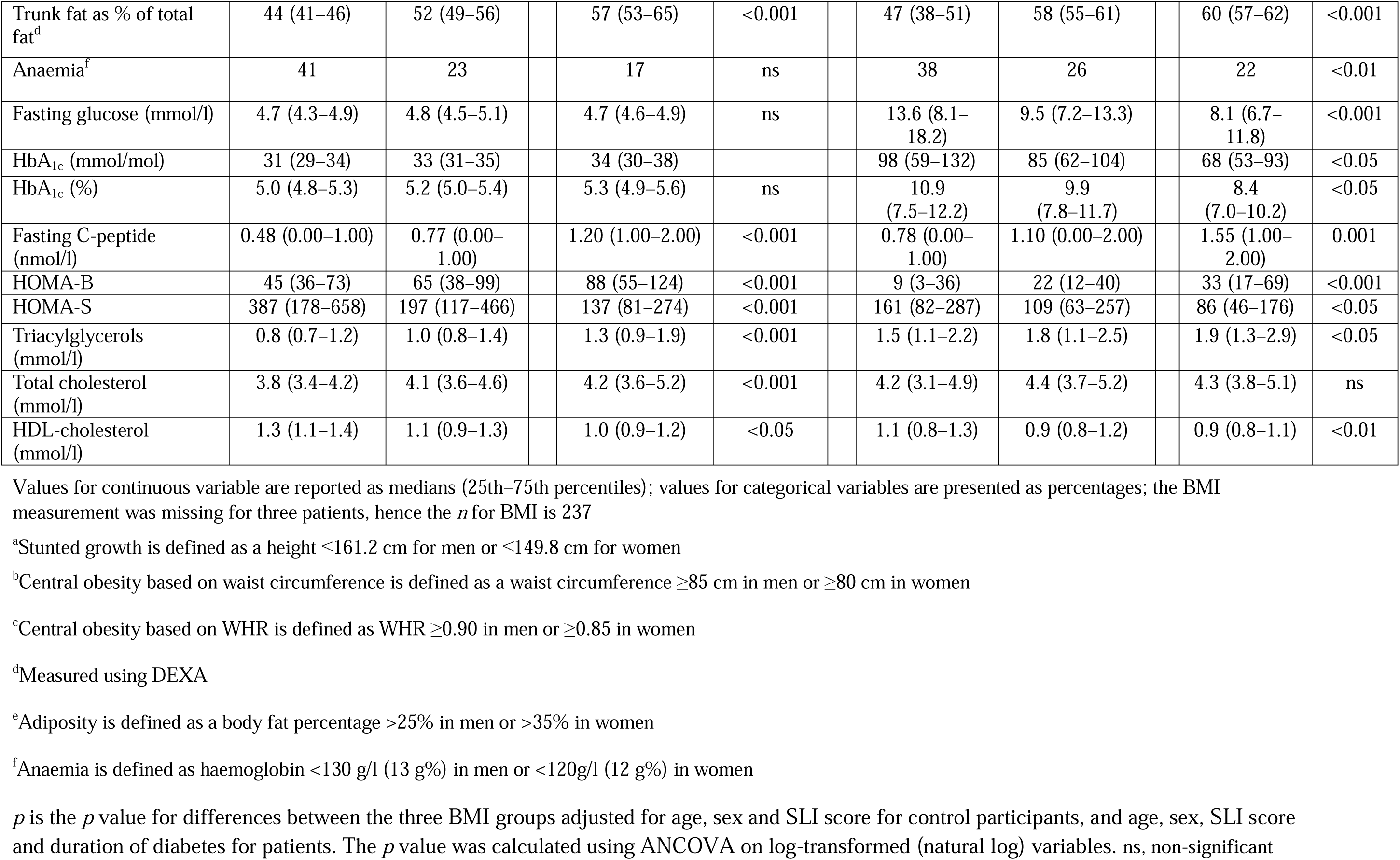
Characteristics of young patients with diabetes and control participants attending the outpatient department of AMCH, according to BMI groups.

### Metabolic endocrine measurements

Most patients with diabetes had suboptimal glucose control, reflected in high glucose and HbA_1c_ concentrations (Table 1); only 40 (17%) had an HbA_1c_ <52 mmol/mol (7.0%). Patients had higher triacylglycerol and lower HDL-cholesterol concentrations compared with control participants; 27% of patients had hypercholesterolaemia (≥5.2 mmol/l) and 58% had hypertriglyceridemia (≥1.7 mmol/l). Low HDL-cholesterol concentrations were observed in 62% of male patients (<1.0 mmol/l) and 76% of female patients (<1.3 mmol/l).

Fasting plasma C-peptide concentrations were low in the diabetic patients: 29% had a concentration <0.5 nmol/l, but none had a history of diabetic ketoacidosis. The HOMA2 estimates of beta cell function and insulin sensitivity relate to healthy UK individuals who are allocated a value of 100 for both measures [36]. Based on these estimates, the control participants with NGT were more insulin-sensitive (HOMA-S value is approximately twice the reference) and had lower beta cell function (HOMA-B value is approximately two-thirds of the reference). Compared with the control participants, the diabetic patients had lower HOMA-B and HOMA-S (Fig. 2). The deficit in HOMA-B was greater (approximately 70% lower on average than in control participants) compared with that in HOMA-S (approximately 50% lower). In Fig. 3, the hyperbolic plots of HOMA-S and HOMA-B highlight that reduction of HOMA-B is the more prominent pathophysiology in diabetic patients compared with control participants.

**Fig. 3.**
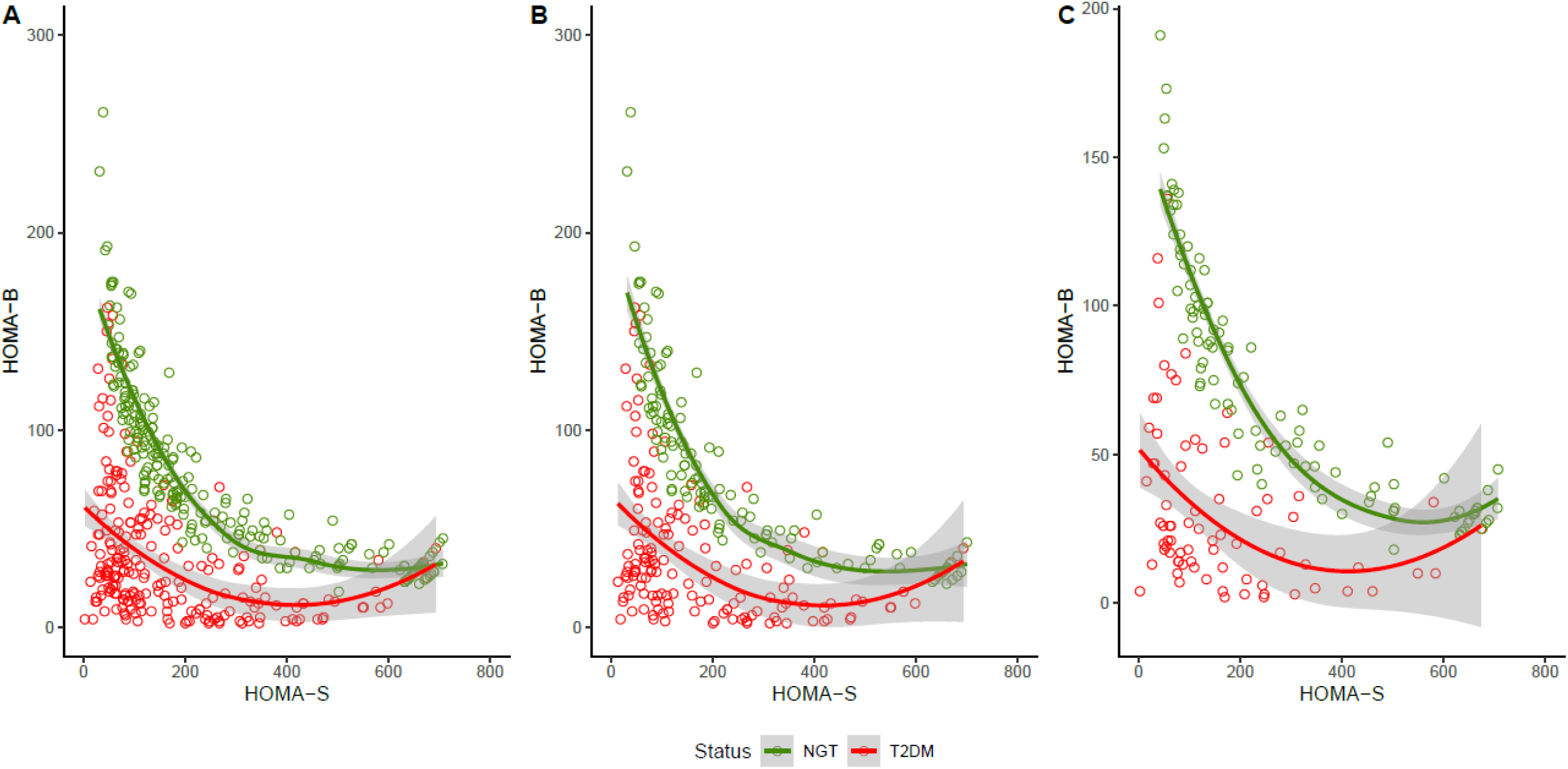
Hyperbolic relationships between HOMA-S and HOMA-B in control participants (green) and young patients with type 2 diabetes (red) for (**a**) all participants, (**b**) male participants and (**c**) female participants. The plots shows that lower HOMA-B is the predominant defect in patients with type 2 diabetes compared with control participants

### Association of BMI with metabolic endocrine parameters

We compared the characteristics of underweight, normal weight, and overweight/obese groups (Table 2). Increasing BMI was associated with higher socioeconomic status (*r*=0.35, *p*<0.001) and urban residence. Underweight patients had the lowest waist circumference, WHR, BRI, adiposity (total and truncal fat percentage assessed using DEXA) and fasting C-peptide and plasma triacylglycerol concentrations, but the highest fasting glucose, HbA_1c_ and HDL-cholesterol concentrations. Underweight patients also had the lowest beta cell function (HOMA-B) and highest insulin sensitivity (HOMA-S). These findings were broadly similar in male participants and female participants. As reported previously [38, 39], the predominant subgroup of type 2 diabetes in our patients, based on the algorithm from Ahlqvist et al, based on a Swedish cohort, was severely insulin-deficient diabetes (SIDD, 67%). ESM Table 3 shows that 77% of patients in the lean BMI category (BMI <25 kg/m^2^) belonged to the SIDD subgroup (79% in the underweight category and 75% in the normal weight category), as did 51% of the overweight/obese patients. Thus, irrespective of the BMI category, the predominant subgroup was SIDD. SIDD was more common in male participants (73% vs 54%), and the mildly obese diabetes (MOD) subgroup was more common in female participants (38% vs 16%).

**Table 3.** Characteristics of age-matched young patients with diabetes and control participants attending the outpatient department of AMCH.

| Characteristics | Male participants (n=142) |  | *p* value |  | Female participants (n=116) |  | *p* value |
| --- | --- | --- | --- | --- | --- | --- | --- |
|  | Control participants (n=71) | Patients (n=71) |  |  | Control participants (n=58) | Patients (n=58) |  |
| Age (years) | 34 (30–36) | 34 (30–36) | ns |  | 33 (30–36) | 33 (30–36) | ns |
| Height (cm) | 166 (162–170) | 165 (162–169) | ns |  | 154 (151–159) | 154 (150–159) | ns |
| BMI (kg/m<sup>2</sup>) | 24 (22–26) | 24 (21–26) | ns |  | 23 (21–25) | 23 (20–26) | ns |
| Waist circumference (cm) | 88 (81–95) | 88 (81–95) | ns |  | 85 (80–90) | 85 (76–94) | ns |
| WHR | 0.95 (0.91–0.98) | 0.96 (0.93–0.99) | ns |  | 0.89 (0.86–0.94) | 0.94 (0.89–0.97) | <0.01 |
| BRI | 9.1 (7.7–10.6) | 9.2 (7.6–10.6) | ns |  | 9.7 (8.8–11.3) | 10.2 (8.1–11.8) | ns |
| Total fat as % of body weight<sup>a</sup> | 29 (24–32) | 28 (23–32) | ns |  | 38 (30–42) | 34 (27–38) | ns |
| Trunk fat as % of total fat<sup>a</sup> | 57 (55–59) | 58 (56–61) | ns |  | 51 (49–54) | 57 (54–60) | <0.001 |
| Fasting glucose (mmol/l) | 4.8 (4.6–5.0) | 8.8 (6.4–12.1) | <0.001 |  | 4.8 (4.7–5.1) | 10.1 (6.4–13.5) | <0.001 |
| HbA<sub>1c</sub> (mmol/mol) | 31 (31–42) | 75 (64–97) | <0.001 |  | 31 (31–31) | 75 (53–105) | <0.001 |
| HbA<sub>1c</sub> (%) | 5.0 (5.0–6.0) | 8.9 (8.0–11.2) | <0.001 |  | 5.0 (5.0–5.0) | 8.9 (7.6–12.3) | <0.001 |
| Fasting C-peptide (nmol/l) | 1.1 (0.0–1.0) | 1.6 (1.0–2.0) | ns |  | 0.9 (0.0–1.0) | 1.5 (1.0–2.0) | <0.01 |
| HOMA-B | 77 (47–108) | 33 (18–57) | <0.001 |  | 74 (32–107) | 27 (13–54) | <0.001 |
| HOMA-S | 167 (98–305) | 68 (48–130) | <0.001 |  | 158 (102–502) | 82 (49–212) | <0.001 |
| Triacylglycerols (mmol/l) | 1.5 (1.0–2.0) | 1.9 (1.2–3.0) | ns |  | 0.9 (0.8–1.3) | 1.8 (1.1–2.5) | <0.001 |
| Total cholesterol (mmol/l) | 4.5 (4.0–5.3) | 4.4 (3.7–5.0) | ns |  | 4.2 (3.6–4.8) | 4.5 (4.1–5.2) | <0.05 |
| HDL-cholesterol (mmol/l) | 1.1 (0.9–1.2) | 0.9 (0.7–1.1) | <0.01 |  | 1.2 (1.0–1.4) | 1.0 (0.8–1.2) | <0.01 |

Values for continuous variable are reported as medians (25th–75th percentiles); values for categorical variables are presented as percentages
The patients and control participants were matched according to age and sex using the ‘matchit’ function in R
<sup>a</sup>Measured using DEXA
*p* values were calculated using the Wilcoxon rank sum test for continuous traits and Fisher’s exact test for categorical traits. ns, non-significant

It is noteworthy that, despite a lower median BMI compared with contemporary non-Hispanic white Americans (23 vs 28 kg/m^2^), our Assamese cohort had higher WHR, BRI and total and truncal adiposity across the range of BMI (including in underweight and lean patients) (Fig. 1).

## Sensitivity analysis

### Age

Given that the diabetic patients were older than control participants, we compared age-matched patients and control participants (Table 3). This generated a dataset of 258 participants (129 patients and 129 control participants, each including 71 male participants and 58 female participants, with an median age of 34 years in male participants and 33 years in female participants). The results are broadly similar to those of the original analysis: the diabetic patients have similar BMI compared with control participants, with higher WHR and triacylglycerols and lower HDL-cholesterol. The diabetic patients also had higher glucose and HbA_1c_, and lower beta cell function and insulin sensitivity.

### Duration of diabetes

The duration of diabetes ranged from <1 year to 19 years. As expected, duration of diabetes was associated with age. Body size and composition, beta cell function and insulin sensitivity were comparable across the duration of diabetes categories when adjusted for age and sex (Table 4). Compared with control participants, newly diagnosed patients (<1 year) had higher WHR and truncal adiposity and lower beta cell function and insulin sensitivity.

**Table 4.** Characteristics of young patients with diabetes attending the outpatient department of AMCH, stratified by duration of diabetes.

|  | Male participants (n=288) |  |  |  |  |  |  | Female participants(n=199) |  |  |  |  |  |
| --- | --- | --- | --- | --- | --- | --- | --- | --- | --- | --- | --- | --- | --- |
| | Control participants (n=136) | Patients (n=152) | | | $p_1$ | $p_2$ | | Control participants (n=116) | Patients (n=83) | | | $p_1$ | $p_2$ |
| Duration of diabetes |  | <1 year (n=72) | 1–5 years (n=46) | >5 years (n=34) |  |  |  |  | <1 year (n=23) | 1–5 years (n=31) | >5 years (n=29) |  |  |
| Age (years) | 29 (25–34) | 34(31–37) | 36 (32–39) | 38 (34–41) | <0.01 | <0.001 |  | 30 (27–35) | 33 (27–36) | 36 (32–40) | 37 (34–40) | <0.05 | ns |
| Height (cm) | 167 (162–172) | 165 (160–169) | 164 (161–168) | 165 (160–170) | ns | <0.05 |  | 155 (151–159) | 155 (152–159) | 153 (149–155) | 153 (150–158) | ns | ns |
| BMI (kg/m<sup>2</sup>) | 23 (21–25) | 24 (21–26) | 23 (20–26) | 23 (19–26) | ns | ns |  | 23 (21–25) | 24 (20–25) | 23 (20–25) | 23 (21–24) | ns | ns |
| Waist circumference (cm) | 84 (78–92) | 88 (80–95) | 86 (78–91) | 89 (77–94) | ns | ns |  | 85 (78–90) | 87 (78–93) | 85 (80–90) | 87 (77–95) | ns | ns |
| WHR | 0.93 (0.88–0.96) | 0.96 (0.92–0.99) | 0.96 (0.91–0.99) | 0.98 (0.92–1.01) | ns | <0.001 |  | 0.9 (0.86–0.94) | 0.94 (0.89–1) | 0.94 (0.89–0.96) | 0.94 (0.9–1) | ns | <0.01 |
| BRI | 8.1 (6.8–9.9) | 8.9 (7.6–10.7) | 8.8 (7.2–9.9) | 9.9 (6.9–10.7) | ns | <0.05 |  | 9.7 (7.8–11.3) | 10.7 (8.1–11.9) | 10.2 (7.9–11.4) | 10.6 (8.3–11.9) | ns | ns |
| Total fat as % of body weight<sup>a</sup> | 28 (20–31) | 28 (18–32) | 25 (19–29) | 25 (22–32) | ns | ns |  | 38 (29–41) | 35 (28–40) | 35 (31–37) | 32 (23–39) | ns | ns |
| Trunk fat as % of total fat<sup>a</sup> | 55 (50–59) | 59 (55–62) | 57 (53–61) | 59 (54–61) | ns | <0.05 |  | 51 (48–54) | 59 (54–60) | 57 (53–61) | 55 (53–59) | ns | <0.001 |
| Fasting glucose (mmol/l) | 4.8 (4.5–5.0) | 8.2 (6.1–12.4) | 8.8 (6.7–11.2) | 11.4 (7.8–13.6) | ns | <0.05 |  | 4.7 (4.5–4.9) | 8.2 (5.2–13.1) | 11.5 (8.1–13.6) | 11.2 (8.2–13.6) | ns | <0.001 |
| HbA<sub>1c</sub> (mmol/mol) | 31 (31–42) | 86 (61–108) | 75 (53–97) | 86 (64–113) |  | <0.001 |  | 31 (31–31) | 64 (45–116) | 86 (61–99) | 86 (64–97) |  | <0.001 |
| HbA<sub>1c</sub> (%) | 5.0 (5.0–6.0) | 10.0 (7.5–12.0) | 8.0 (6.8–10.9) | 10.0 (7.9–12.5) | ns | <0.001 |  | 5.0 (5.0–5.0) | 8.1 (6.3–12.8) | 10.0 (7.5–11.2) | 10.0 (7.9–12.5) | ns | <0.001 |
| Fasting C-peptide (nmol/l) | 0.97 (0.00–1.00) | 1.10 (0.00–2.00) | 1.10 (0.00–2.00) | 1.30 (1.00–2.00) | ns | ns |  | 0.78 (0.00–1.00) | 1.30 (0.00–2.00) | 1.50 (1.00–2.00) | 1.10 (0.00–2.00) | ns | <0.05 |
| HOMA-B | 69 (43–108) | 30 (13–53) | 30 (12–47) | 22 (12–31) | ns | <0.001 |  | 65 (33–99) | 29 (16–55) | 23 (11–52) | 21 (13–30) | ns | <0.001 |
| HOMA-S | 175 (98–353) | 117 (63–319) | 103 (64–217) | 81 (49–187) | ns | <0.05 |  | 201 (115–502) | 104 (50–288) | 81 (61–150) | 115 (58–243) | <0.05 | <0.05 |
| Triacylglycerols (mmol/l) | 1.2 (0.9–1.9) | 1.0 (0.8–1.3) | 1.8 (1.2–2.5) | 1.6 (1.1–2.3) | ns | <0.01 |  | 1.0 (0.8–1.3) | 1.8 (1.2–2.5) | 1.6 (1.1–2.3) | 2.0 (1.4–3.3) | ns | <0.001 |
| Total cholesterol (mmol/l) | 4.1 (3.6–4.7) | 4.1 (3.5–4.7) | 4.3 (3.6–5.1) | 4.2 (3.5–4.5) | ns | ns |  | 4.1 (3.5–4.7) | 4.3 (3.6–5.1) | 4.2 (3.5–4.5) | 4.7 (4.1–5.5) | ns | <0.05 |
| HDL-cholesterol (mmol/l) | 1.1 (0.9–1.2) | 1.2 (1.0–1.4) | 1.0 (0.8–1.2) | 0.9 (0.8–1.1) | ns | ns |  | 1.2 (1.0–1.4) | 1.0 (0.8–1.2) | 0.9 (0.8–1.1) | 0.9 (0.8–1.1) | ns | <0.001 |

Values for continuous variable are reported as medians (25th–75th percentiles); values for categorical variables are presented as percentages; *n* for patients reflects the number of participants for whom duration of diabetes was available
<sup>a</sup>Measured using DEXA
*p* values were calculated using ANCOVA, adjusted for age and SLI score. *p*<sub>1</sub> is the *p* value for the difference between the three duration groups, *p*<sub>2</sub> is the *p* value for the difference between newly diagnosed patients (<1 year) and control participants. ns, non-significant

## Discussion

We describe the sociodemographic history, and physical and metabolic endocrine characteristics of GADA-negative young patients with diabetes among impoverished tribal tea garden workers in Assam in North-East India. Our comprehensive description of body size, shape and composition (based on DEXA measurements) and metabolic endocrine features defines a unique phenotype of young people with diabetes in this population. There was a high prevalence of stunting and anaemia, highlighting multigenerational socioecological deprivation and undernutrition. Interestingly, this was coupled with higher central obesity/adiposity (WHR, BRI and truncal adiposity) compared with control participants as well as contemporary non-Hispanic white Americans in the NHANES cohort, supporting the existence of the thrifty thin–fat phenotype. Our research has shown that such a thin–fat phenotype is programmed by *in utero* undernutrition, and continues into childhood, puberty and adult life [6–10,40]. Diabetes in this population was predominantly due to reduced beta cell function, with only a moderate contribution from reduced insulin sensitivity. Although relatively lean and beta cell-deficient, these patients had higher triacylglycerol and lower HDL-cholesterol concentrations compared with control participants, reflecting a probable contribution of hepatic adiposity. Intriguingly, potbelly, fatty liver and shrunken pancreas have been described in undernourished children in India [41] and other countries [42], and those findings support the hypothesis that early-life undernutrition contributes to this phenotype. The Developmental Origins of Health and Disease (DOHaD) paradigm proposes that non-communicable diseases are programmed epigenetically during the periconceptional period by the interaction between genes and the intrauterine environment [43]. Assamese mothers have a higher prevalence of underweight (25.9%) compared with the Indian national average (22.9%), and there is a high prevalence of low birthweight and undernutrition in children under 5 years old in this population [44]. It is possible that uncontrolled long-standing metabolic disturbances (glucotoxicity and lipotoxicity) made a contribution to low beta cell function (insulin deficiency) in these patients. However, the characteristic body phenotype and lower beta cell function (in relation to the reference population of the HOMA2 model [36]) even in the control participants suggest that these are intrinsic characteristics of this chronically undernourished population. The parents and grandparents of the majority of participants were exposed to the Bengal famine in 1943 as children or young adults, and also to colonial harshness and slavery. The population of North-East India has also been exposed to socioeconomic deprivation and political unrest over long periods of time (M. Thakur [Ministry of Tribal Affairs, Government of India], personal communication, 15 January 2025, reproduced as ESM Background Text), providing a background to the multigenerational origin of the phenotypes in our study population.

Thus, the physical, nutritional and metabolic endocrine characteristics of these diabetic patients differ substantially from the conventional descriptions of young, obese and insulin-resistant type 2 diabetes patients from the Western world [45], urban India [12] and Indigenous populations from various parts of the world [46–48]. A direct comparison with Swedish young-onset type 2 diabetes patients using the Swedish clustering algorithm revealed that two-thirds of our patients from Assam belonged to the SIDD subgroup and a quarter belonged to the MOD subgroup, which is the opposite of the proportions in Swedish patients (one-quarter SIDD; two-thirds MOD) [39]. Although the SIDD subgroup was predominant across the BMI groups, the decreasing proportion of SIDD and the increasing insulin resistance with increasing BMI highlight the evolution of diabetes phenotypes with urbanisation and socioeconomic transition in this population. Indeed, a third of these patients were overweight/obese, demonstrating the double burden of malnutrition. Thus, our results provide a snapshot of the evolution of the phenotype of young non-autoimmune diabetes in transitioning populations. It is interesting to note that intrauterine undernutrition programmed future obesity in the Dutch Hunger Winter [49] if the mother was exposed to famine in the first two trimesters of pregnancy. In addition, it has been shown that multigenerationally undernourished rats become obese and hyperinsulinaemic within two generations of ‘normal’ feeding [50]. It is noteworthy that early-life undernutrition is also linked with the risk of diabetes in the contemporary NHANES cohort in the USA: low birthweight increased the risk of prediabetes in adolescents (ADA criteria) [51], and stunting increased the risk of diabetes in adults [52, 53]. Despite such evidence, attention is usually focused on later-life obesity as the sole preventable risk factor for diabetes, thus missing the crucial role of early-life undernutrition, which could inform strategies for primordial prevention. To the best of our knowledge, ours is the largest and most detailed report of young-onset, non-autoimmune diabetes in North-East India. Measurement of immunoreactive trypsinogen levels helped to exclude exocrine pancreatic diabetes. Detailed anthropometric and DEXA body composition measurements, epidemiological indices of beta cell function and insulin sensitivity (C-peptide-based HOMA) helped us to characterise these patients better than many past reports from India. Admittedly, it is a hospital-based study, but it has helped to define the unique phenotype of diabetes in a deprived population and search for mechanistic explanations. Our data also show interesting differences between male and female patients: SIDD was more common in male patients and MOD was more common in female patients, suggesting possible aetiological sex differences and also providing useful guidelines for treatment options. Our findings may apply to impoverished patients with diabetes in other low– and middle-income countries. This is evident in the similarity in the phenotype of our patients compared with patients with non-autoimmune young-onset diabetes from Uganda, Ethiopia, Jamaica and rural Vellore, Tamil Nadu (India) [17–20, 23–24]. The Vellore study [17] helped to validate the findings of epidemiological measurements (HOMA-S and HOMA-B) using state-of-the-art techniques. Despite differences in genetic and sociocultural factors, and the methodology of these studies, all the patients were lean, beta cell-deficient and insulin-sensitive. This suggests the existence of a unique subgroup of non-autoimmune young-onset diabetes in impoverished populations. An international workshop in India recently proposed that this phenotype be called ‘type 5 diabetes’, and encouraged further research on the subject [54]. The cross-sectional design of our study precludes attribution of causality to undernutrition; however, such causality is supported by prospective metabolic endocrine observations in people with severe acute childhood malnutrition in Jamaica [23,24], animal models [50] and birth cohort research in India [55]. The Pune Maternal Nutrition Study (which studied a rural Indian birth cohort) showed that prediabetes (impaired fasting glucose and impaired glucose tolerance) in young and lean participants is linked to intrauterine growth restriction (stunting and smaller head circumference at birth) and lower compensatory insulin secretion (disposition index) from early childhood [55]. There remains a need for longitudinal studies in North-East India. A study of body composition and glucose/insulin metabolism in children born in AMCH has recently been completed, and it is hoped that this will develop into a longitudinal cohort (ICMR grant number 55/3/2/Brain Storming Sessions/diab/2017-NCD-II).

In conclusion, we report the characteristics of young-onset, non-autoimmune patients with diabetes in the impoverished population of Assam, North-East India. These patients have low lean mass and excess central and generalised adiposity at a relatively low BMI compared with non-diabetic control participants and white Americans. Their metabolic profile includes dominant insulin secretory failure, relatively preserved insulin sensitivity and characteristic dyslipidaemia. Such a metabolic profile would predispose these patients to micro– and macrovascular complications at the peak of their productive lives. The implications are far-reaching, both for the individual patients and for the resource-constrained health system. Our data support devising health strategies to improve life-course nutrition to help prevent diabetes (primordial prevention), and research into and customisation of diabetes treatment to reduce morbidity and mortality, rather than following Western guidelines. Prospective longitudinal life-course studies of genetic and epigenetic factors, nutrition and growth will improve our understanding of the aetiological factors of diabetes in this population and guide prevention. Our findings emphasise the important contribution of social determinants of health in the heterogeneity of type 2 diabetes phenotypes across the world, and stress the need to address the relevant sustainable development goals (poverty, hunger and equality; https://sdgs.un.org/goals) to prevent and manage diabetes in these patients. These issues are becoming increasingly relevant in food-insecure populations affected by wars and conflict, displacement and climate change.

## Supporting information

This is the ESM file

## Acknowledgements

We thank all patients and control participants for their support and willingness to participate. We thank H. Kashyap, H. Kaur, A. Gogoi, A. Boruah, D. Boruah, S. Sonowal, P. Boruah, R. Nath, I. Gogoi, P. Gogoi, D. Ahmed (Assam Medical College and Hospital), P. Yajnik and R. Ladkat (Diabetes Unit, Kamalnayan Bajaj Diabetology Research Centre, King Edward Memorial Hospital and Research Centre, Pune, India) and L. Saikia and R. Nath (Multidisciplinary Research Unit, Assam Medical College and Hospital) for their invaluable contributions to this study. We profusely thank M. Thakur, Ministry of Tribal Affairs, Government of India, for providing the socioeconomic background document. We thank N. Gupte, Johns Hopkins India Private Limited, Mumbai, India, for advice on statistical analysis.

## Abbreviations

AMCH: Assam Medical College and Hospital
BRI: Body roundness index
DEXA: Dual-energy x-ray absorptiometry
MOD: Mildly obese diabetes
NGT: Normal glucose tolerance
PHENOEINDY-2: PHEnotypingNOrthEastINDianYoung type 2 diabetes
SIDD: Severely insulin-deficient diabetes

## Data availability

The datasets generated during and/or analysed during the current study are available from the corresponding author on reasonable request, subject to necessary administrative permission.

## Authors’ relationships and activities

The authors declare that there are no relationships or activities that might bias, or be perceived to bias, their work.

## Contribution statement

AD, CSY and SK contributed to the conception of the work. AD, PKD, PD, SMB and AR contributed to data collection. DSB, SWP, MD, RM, TL, PT and RW contributed to the data analysis. CSY, AD, RM, TL and RW drafted the article. All authors contributed to the interpretation of data and critical revision of the article. All authors gave final approval of the version to be published. AD, CSY and SK are the guarantors of this work, and, as such, had full access to all the data in the study and take responsibility for the integrity of the data and the accuracy of the data analysis.

