## Supplementary material for "Non-autoimmune diabetes in young people from Assam, India: the PHENOEINDY-2 study": This is the ESM file

**ELETRONIC SUPPLEMENTARY MATERIAL**

**ESM Text (background) 1: Colonialism, Identity Conflict, and Poverty in Northeast India: Historical Roots and Modern Struggles**

Northeast India has a long history of violence, exploitation, and economic underdevelopment despite its strategic location and great cultural diversity. The region has seen a complicated interplay of identity problems, alienation, and resistance since colonial times to the present, which has been made worse by economic marginalization. This article looks at how indigenous groups like the Kukis, Mizos, and Assamese tea tribes were altered by British colonialism and how these historical upheavals have fuelled militancy and poverty in Assam, especially with the emergence of the United Liberation Front of Assam (ULFA). The cycles of poverty and violence that still exist in the area now can be better understood by looking at the historical causes of identity-based conflict and economic exclusion.

**Colonial Penetration and Indigenous Resistance**

Indigenous populations' social, economic, and cultural dynamics were changed by the British colonial presence in Northeast India. The 19th-century Kuki resistance is one prominent example. Kuki incursions on colonial outposts were frequently depicted by British officials as acts of lawlessness. In actuality, though, these raids were a kind of coordinated opposition to the British invasion of Kuki land and their attempts to impose colonial laws and political systems. Like many other indigenous communities in the area, the Kukis had developed governance structures and territory claims that ran counter to British imperial aspirations. The colonial state's attempts to erode their political independence and cultural customs were categorically rejected by their resistance [1].

Similarly, the colonial encounter had a significant impact on Mizo civilization in the Lushai Hills. The Mizos engaged in jhum, or shifting cultivation, prior to British interference; this technique was strongly related to their cultural and environmental customs. In order to harvest resources like timber for the British empire, colonial policies attempted to control land and forest use, not for the benefit of the native population. The colonial state and Mizo chiefs, who had traditionally been in charge of allocating land within their villages, became at odds when these policies were imposed, disrupting traditional Mizo land-use patterns. The Mizo relationship with the environment was drastically changed by the British as they imposed scientific rationalism and economic goals, leading to new conflicts that would have a lasting impact [2,3].

Christian missionaries were also active during the colonial era and were instrumental in the sociocultural change of native communities. Welsh missionaries like Dr. Peter Fraser fought the colonial authorities in the Lushai Hills over the Bawi system, which was a Mizo system of enslavement. Fraser advocated for the eradication of the Bawi system because he saw it as a kind of slavery. Fraser's attempts were thwarted by the colonial government, which was reluctant to disturb the tenuous peace it had made with the Mizo chiefs. This conflict between the missionaries and the colonial authority demonstrates the larger conflicts between the missionaries' attempt to force Western morality and modernity on indigenous populations and the desire to preserve control and order [3].

**Exploitation of the Tea Tribes in Assam**

In the Assamese tea industry, where British planters depended on the forced migration and coercion of Adivasi workers from areas like Jharkhand, Odisha, and Chhattisgarh, colonial exploitation took on its most obvious form. These workers, referred to as the "tea tribes," were transported to Assam and forced to work in substandard conditions in ghettoized "coolie lines" with limited mobility. By enforcing harsh labor regulations like the Workmen's Breach of Contract Act, which essentially bound workers to the estates, British colonial administration aided the planters. Despite making substantial contributions to Assam's tea economy, these labourers continued to be economically and socially disenfranchised. Their exploitation reflected the larger colonial strategy of labor enslavement and resource extraction, producing a legacy of exclusion and hardship that still affects Assam's socioeconomic situation [4].

**Militancy, Poverty, and Identity Conflict in Assam**

Assam's ongoing resource and identity problems derive from the colonial history and neglect after independence. The United Liberation Front of Assam (ULFA), created in 1979, sought to free Assam from central rule, motivated by long-standing grievances including as poverty, unemployment, and ethnic nationalism. Violence, extortion, and human rights violations became commonplace, resulting in about 30,000 deaths during the 1980s and 1990s. Extrajudicial killings, particularly from 1996 to 2001, fuelled the conflict. Scholars such as Suresh Tendulkar and K. Sundaram have identified a direct link between increased poverty and militancy, particularly from the 1980s to the early 2000s [5].

Economic imbalances between urban and rural areas, worsened by illegal immigration from Bangladesh, spurred ethnic conflict and rivalry for resources. Poverty intensified, with Assam's Below Poverty Line (BPL) population increasing by 1.3 million between the 1980s and 1990s to 31.98% by 2011-12, greatly above the national average of 21.92%. Many young people have become militant as a result of their economic marginalization [5].

The cycle of poverty and insurgency has hampered investment and growth, further isolating Assam. Repressive policies, such as the Armed Forces Special Powers Act (AFSPA), have exacerbated alienation among locals. Recently, government initiatives to reduce poverty through infrastructure development, education, and job creation, as well as calls for engagement with ULFA, have aimed to break the region's long-standing cycle of poverty and violence [5].

**Conclusion**

In Assam and the larger Northeast, colonialism left a long-lasting legacy of turmoil, isolation, and exploitation. The colonial state changed indigenous society in ways that still have an impact on the area today, from the exploitation of tea workers to the Kuki resistance and Mizo environmental issues. These issues have been made worse by post-colonial practices of economic neglect and exclusion, which have resulted in Assamese poverty and militancy cycles. Assam's poverty greatly increased between 1983 and 2012, which fuelled the insurgency and added to the violence that resulted in loss of unprecedented number of lives. Going forward, the best way to end these cycles and promote long-term peace and prosperity for the area is to take a comprehensive approach that incorporates development, dialogue, and inclusive policy.

**ESM Table 1: Socio-Demographics of the young cases with diabetes and controls attending AMCH, Dibrugarh, Assam**

| **Characteristics** | **Overall (*N=492*)** | **Cases (*N=240*)** | **Controls (*N=252*)** | ***p-value*** |
| --- | --- | --- | --- | --- |
| Gender  Men  Women | 291 (59%)  201 (41%) | 155 (64.6%)  85 (35.4%) | 136 (54%)  116 (46%) | **0.011** |
| Linguistic group  Assamese  Bengali  Others | 297 (60%)  79 (16%)  116 (24%) | 150 (62.5%)  45 (18.8%)  45 (18.7%) | 147 (58%)  34 (14%)  71 (28%) | **0.029** |
| Religion  Hindu  Muslim  Others | 418 (85%)  68 (14%)  6 (1%) | 198 (82.5%)  40 (16.7%)  2 (0.8%) | 220 (87%)  28 (11%)  4 (2%) | 0.190 |
| Physical activity (N=489)  Sedentary  Light active  Moderate to highly active | 129 (26%)  227 (46%)  130 (27%) | 70 (29.5%)  109 (46.0%)  58 (24.5%) | 59 (23%)  118 (47%)  72 (30%) | 0.263 |
| Smoking | 78 (16%) | 33 (13.7%) | 45 (18%) | 0.173 |
| Pan consumption | 286 (58%) | 135 (56%) | 151 (60%) | 0.496 |
| Alcohol consumption | 145 (30%) | 61 (25.4%) | 84 (33%) | 0.092 |
| Alcohol consumption > thrice a week | 66 (46%) | 31 (52%) | 35 (42%) | 0.023 |

- Values reported as n (%);
- *p-value* calculated using χ^2^ tests

**ESM Table 2: DEXA Measurements of the young cases with diabetes and controls attending AMCH, Dibrugarh, Assam**

|  | **Male** | |  | **Female** | |  |
| --- | --- | --- | --- | --- | --- | --- |
|  | **NGT** | **T2DM** | **p-value** | **NGT** | **T2DM** | **p-value** |
| **Fat mass distribution** |  |  |  |  |  |  |
| Arms(kg) | 1.7 (1.3-2) | 1.7 (1.3-2.1) | 0.968 | 2.4 (1.9-2.7) | 2.0 (1.6-2.6) | 0.051 |
| Legs(kg) | 4.8 (3.9-6.1) | 4.3 (3.2-5.6) | 0.018 | 6.8 (5.6-8.2) | 5.0 (4-6.1) | 0.000 |
| Trunk(kg) | 9.9 (6.6-12.9) | 10.5 (6.8-13.1) | 0.415 | 10.8 (8.6-13.2) | 11.2 (9-13.5) | 0.670 |
| Android(kg) | 1.5 (1-2.1) | 1.6 (1-2.2) | 0.186 | 1.7 (1.1-2) | 1.7 (1.3-2.3) | 0.136 |
| Gynoid(kg) | 2.4 (1.9-3) | 2.1 (1.5-2.8) | 0.050 | 3.6 (2.9-4.1) | 2.8 (2.2-3.2) | 0.000 |
| **Lean mass distribution** |  |  |  |  |  |  |
| Arms(kg) | 5.5 (4.9-6.1) | 5.1 (4.4-5.7) | 0.000 | 3.2 (2.9-3.8) | 3.4 (2.9-4.1) | 0.312 |
| Legs(kg) | 14.3 (12.7-16.4) | 13.9 (12.1-15.5) | 0.065 | 10.5 (9.4-11.6) | 9.8 (8.7-11.4) | 0.264 |
| Trunk(kg) | 20.4 (19.1-22) | 21 (19.1-23.5) | 0.137 | 15.8 (13.9-17.1) | 16.4 (15.1-18.3) | 0.001 |
| Android(kg) | 2.9 (2.6-3.1) | 3.0 (2.7-3.4) | 0.013 | 2.2 (2-2.4) | 2.4 (2.2-2.7) | 0.000 |
| Gynoid(kg) | 6.4 (5.9-7.1) | 6.2 (5.5-6.8) | 0.016 | 4.8 (4.3-5.2) | 4.5 (4.2-5.1) | 0.555 |

- Values are presented as median (25^th^-75^th^ centiles).
- p-values are calculated by *t*-test adjusted for age and standard of living index (SLI) score.

**ESM Table 3: Proportion of subgroups of type 2 diabetes within the BMI groups and within sexes**

|  | **Severe insulin deficiency diabetes (SIDD)** | **Severe insulin resistance diabetes (SIRD)** | **Moderately obese diabetes**  **(MOD)** | **Mild age-related diabetes**  **(MARD)** | **Total** |
| --- | --- | --- | --- | --- | --- |
| Underweight  (<18.5 kg/m^2^) | 79% | 4% | 4% | 13% | 100 % |
| Normal  (18.5-24.9 kg/m^2^) | 75% | 1% | 14% | 10% | 100 % |
| Overweight/Obese (≥25.0 kg/m^2^) | 51% | 0% | 46% | 3% | 100 % |
| Total | 67% | 1% | 24% | 8% | 100 % |
| Male | 73% | 1% | 16% | 10 % | 100% |
| Female | 54% | 4% | 38% | 4% | 100% |

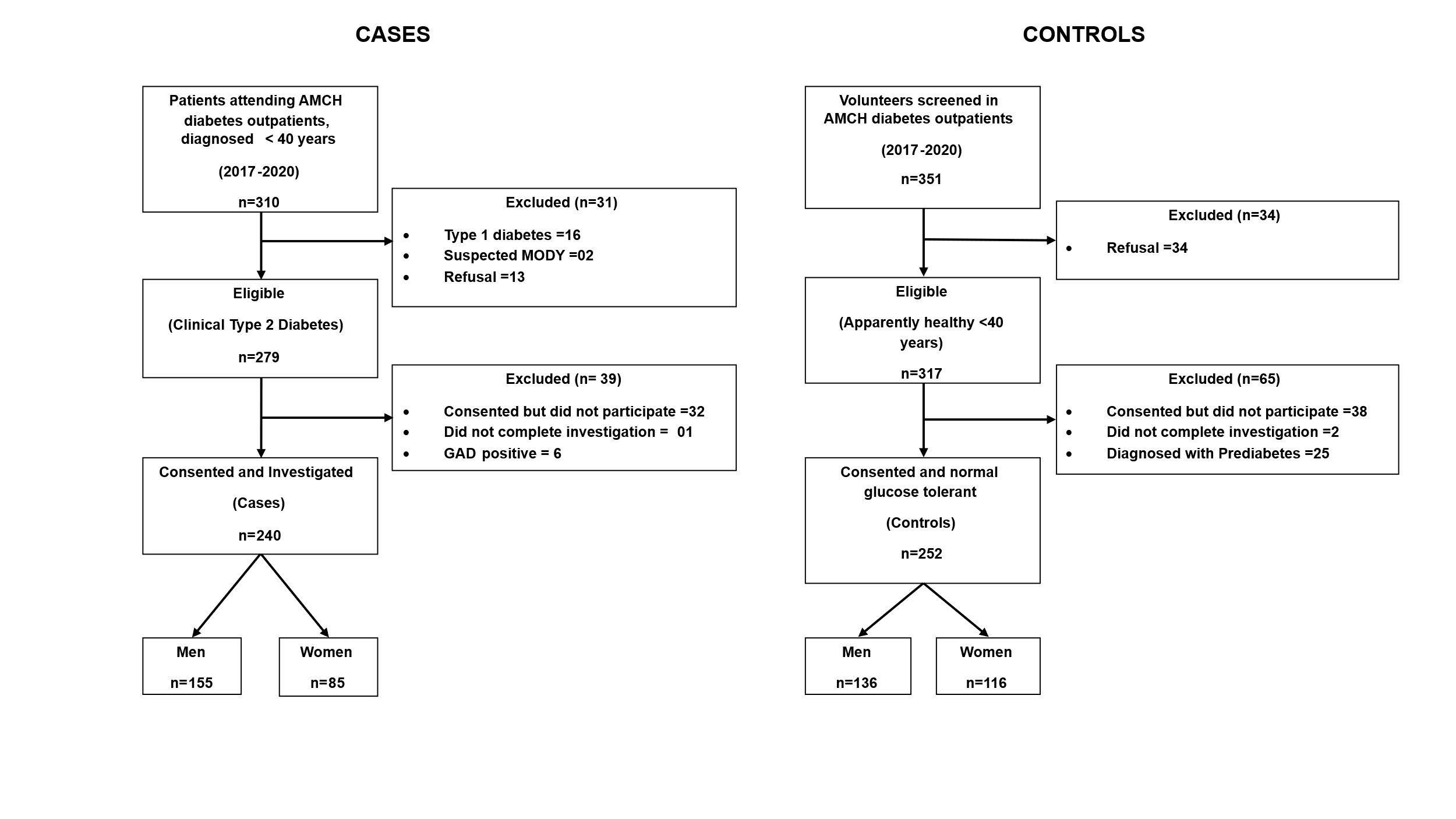

**ESM Figure 1- Flowchart depicting participant selection and flow in the study**

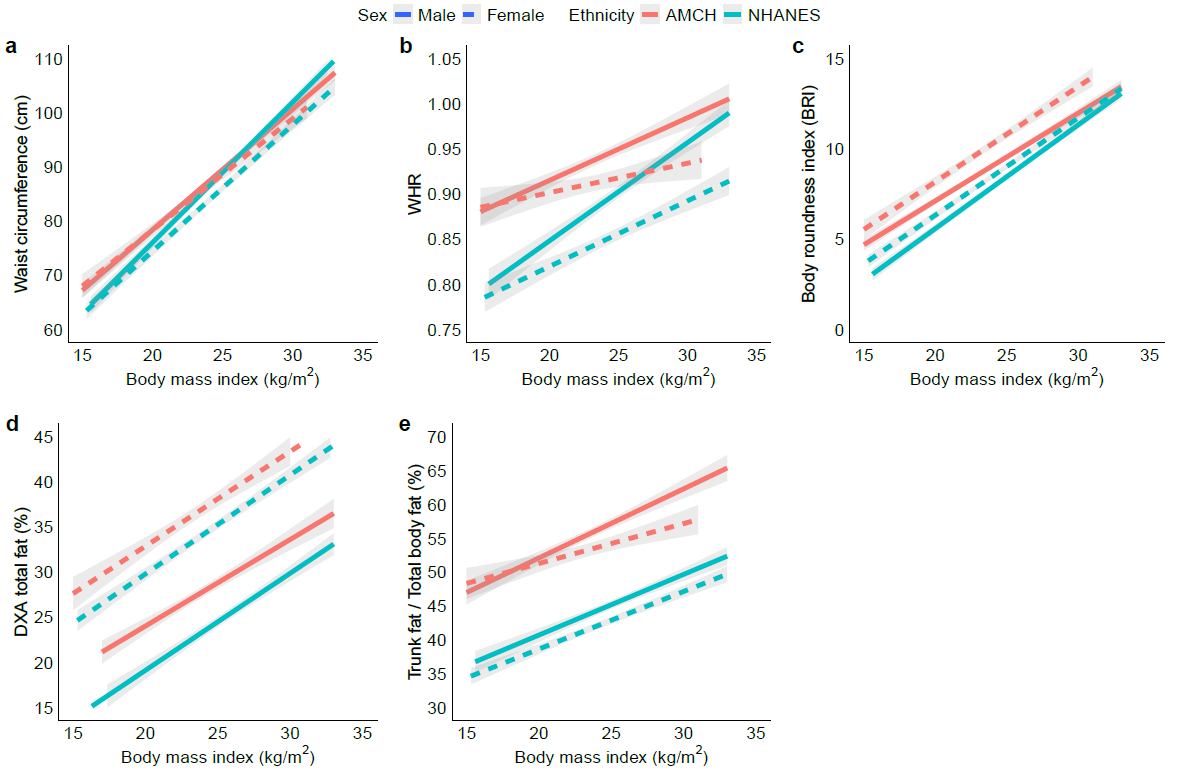

**ESM Figure 2:** Figure shows comparison of waist circumference, WHR, body roundness index (BRI), DXA total and truncal adiposity in participants from Assam Medical College (red line) and non-Hispanic white Americans (NHANES 2017-20) separately for sexes – Males (solid line), Females (dotted line).
